# Trauma informed co-production: Collaborating and combining expertise to improve access to primary care with women with complex needs

**DOI:** 10.1101/2023.04.21.23288783

**Authors:** Helen McGeown, Lucy Potter, Tracey Stone, Julie Swede, Helen Cramer, Bridging Gaps group, Jeremy Horwood, Maria Carvalho, Florrie Connell, Gene Feder, Michelle Farr

**Affiliations:** Centre for Academic Primary Care, Population Health Sciences, Bristol Medical School, University of Bristol, Canynge Hall, 39 Whatley Road, Bristol, BS8 2PS; The National Institute for Health and Care Research Applied Research Collaboration West (NIHR ARC West) at University Hospitals Bristol and Weston NHS Foundation Trust, UK

## Abstract

**Introduction:** Health, social care, charitable and justice sectors are increasingly recognising the need for trauma-informed services that seek to recognise signs of trauma, provide appropriate paths to recovery, and ensure that services enable people rather than re-traumatise. Foundational to the development of trauma-informed services is collaboration with people with lived experience of trauma. Co-production principles may provide a useful framework for this collaboration, due to their emphasis on lived experience, and intent to address power imbalances and promote equity. This article aims to examine trauma-informed and co-production principles to consider the extent to which they overlap and explore how to tailor co-production approaches to support people who have experienced trauma.

**Methods:** Bridging Gaps is a collaboration between women who have experienced complex trauma, a charity that supports them, primary care clinicians and health researchers to improve access to trauma-informed primary care. Using co-production principles, we aimed to ensure that women who have experienced trauma were key decision-makers throughout the project. Through reflective notes (n=19), observations of meetings (n=3), interviews with people involved in the project (n=9) and reflective group discussions on our experiences, we share learning, successes and failures. Data analysis followed a framework approach, using trauma-informed principles.

**Results:** Co-production processes can require adaptation when working with people who have experienced trauma. We emphasise the need for close partnership working, flexibility, and transparency around power dynamics, paying particular attention to aspects of power that are less readily visible. Sharing experiences can re-trigger trauma. People conducting co-production work need to understand trauma and how this may impact upon an individual’s sense of psychological safety. Long-term funding is vital to enable projects to have enough time for establishment of trust and delivery of tangible results.

**Conclusions:** Co-production principles are highly suitable when developing trauma-informed services. Greater consideration needs to be given as to whether and how people share lived experiences, the need for safe spaces, honesty and humility, difficult dynamics between empowerment and safety, and whether and when blurring boundaries may be helpful. Our findings have applicability to policy-making, funding and service provision to enable co-production processes to become more trauma-informed.

**Public Contribution:** Bridging Gaps was started by a group of women who have experienced complex trauma, including addiction, homelessness, mental health problems, sexual exploitation, domestic and sexual violence, and poverty, with a GP who provides healthcare to this population, alongside a support worker from the charity, One25, a charity that supports some of the most marginalised women in Bristol to heal and thrive. More GPs and healthcare researchers joined the group and they have been meeting fortnightly for a period of four years with the aim of improving access to trauma-informed primary care. The group uses co-production principles to work together, and we aim to ensure that women who have experienced trauma are key decision-makers throughout our work together. This article is a summary of our learning, informed by discussion, observations and interviews with members of the group.

## Introduction

Co-production is an approach where professionals and people who may use or are affected by public services collaborate, using the experience and expertise of all to form equitable partnerships to develop services/ research and outcomes^1-3^. Whilst co-production can help to tackle inequities in health services by enabling those who are often excluded to help shape them^1^, there can be barriers that prevent involvement and perpetuate marginalisation^1, 4, 5^. There is increasing recognition of the needs of marginalised communities and the structural obstacles to involvement that they may face^6^, alongside support that might be needed by those who have experienced trauma^7, 8^. Existing co-production approaches need to acknowledge and understand trauma and its potential impact on individuals, group dynamics and health inequalities^8^. The high prevalence of trauma in all sectors of society, and even higher prevalence amongst groups who experience health inequalities, mandates careful consideration of the impact of trauma within co-production work.

Trauma can be defined as “an event,… or set of circumstances that is experienced by an individual as physically or emotionally harmful or life threatening”^9^. Complex trauma is prolonged, and often inflicted by an individual who should be trusted – e.g. experience of child abuse or domestic violence^10^. The reduction in life expectancy for those experiencing complex trauma is well documented^11, 12^, with higher rates of mental health problems, substance misuse, cardiovascular disease, diabetes, gastrointestinal disorders, and cancer, amongst other conditions^13^. Complex trauma negatively impacts the ability to access healthcare services, experience of these services and ability to participate in research^9^. This compounds the existing direct physical and mental health impacts of complex trauma.

In recent years, there has been a move towards trauma-informed approaches to healthcare delivery^9, 14, 15^. An organisation/ system that is trauma-informed:

> “realises the widespread impact of trauma and understands potential paths for recovery; recognises the signs and symptoms of trauma in clients, families, staff and others involved within the system; and responds by fully integrating knowledge about trauma into policies, procedures and practices, and seeks to actively resist re-traumatisation”^9^ (p.9).

Inherent to a trauma-informed approach is the explicit consideration of the cultural, historical and gender factors affecting health, and redistribution of power in decision-making^9^. Although involvement of those with lived experience of trauma is a core aspect of this approach and some research has highlighted the potential for co-production approaches to contribute to this^16^, there is no trauma-informed framework to guide collaborative processes between people with lived experience and other stakeholders (e.g. researchers, clinicians and managers).

In this article we describe how we used co-production principles in the collaborative development of a trauma-informed primary healthcare intervention called Bridging Gaps. We start by reviewing trauma-informed and co-production principles, highlighting overlap as well as key differences. We then provide further details and aims of the Bridging Gaps project. We illustrate our co-production process to date, which involved using the principles of trauma-informed approaches as codes to analyse participant interviews. Findings are used to provide recommendations to those seeking to adopt co-production methods to work with people who have experienced trauma.

### Comparative analysis of co-production principles and trauma informed principles

Whilst co-production has been described as a ‘vague’ concept, encompassing a range of different collaborative approaches^17^, this article draws on the principles of co-production from highly cited definitional material^2, 18-20^ that recognises the vital role of expertise from lived experience in developing services and research. Key principles of co-production include: sharing power and decision making; adequate resources and shared ownership; equality; diversity of voice; accessibility and reciprocity; valuing all perspectives; and an appreciation of different knowledge and skills^2^. Principles of co-production substantially overlap with trauma-informed approaches. Collaboration with marginalised and less privileged communities is frequently a feature of trauma-informed approaches, which may provide crucial opportunities for building self-efficacy, confidence, skills and worth^7^. An emphasis on changing existing power dynamics through collaboration with and empowerment of individuals with lived experience of trauma is a key aspect of trauma-informed approaches. The six principles of trauma-informed approaches are listed in Table 1 and are compared to the principles underlying co-production approaches.

**Table 1.** Comparison of principles of co-production and trauma-informed approaches

| **Principles of trauma-informed approaches<sup>9, 21</sup>** | **Co-production principles<sup>2, 18-20</sup>** |
| --- | --- |
| **Cultural, Historical and Gender issues**<br>Groups will seek to meaningfully engage with historic, gender and cultural issues in a way that avoids biases and stereotypes. They are aware of and respond to the needs of individuals from different racial, ethnic, religious and cultural backgrounds and different genders. Historical trauma is recognised and addressed, and the potential therapeutic value of cultural connections and gender-specific services is harnessed<sup>9</sup>. | **Accessibility, diversity and inclusion - Respecting and valuing the knowledge of all**<br>Covered by principles such as diversity and inclusion in social care practice literature<sup>19</sup>. Sometimes less explicitly addressed in some co-produced research literature<sup>3</sup>, but more in associated literatures on participatory action research and user-controlled research<sup>22, 23</sup>. Different co-productive methods can be rooted in different critical theory including Marxism, anarchism, critical race theories, feminism, disability rights and indigenous knowledge<sup>17, 24, 25</sup>, which highlight various structural inequalities and acknowledge how power is unequally distributed, creating boundaries and limits of co-production<sup>26, 27</sup>. Not all co-production literature is rooted in these political approaches and there are more instrumental/ tokenistic usages of the concept, which have been critiqued<sup>1, 28</sup>. |
| **Peer Support**<br>Peer support involves people with lived experience of trauma supporting each other directly. This can help some people process their trauma as they share difficult experiences in a supportive environment with other people who understand them<sup>9</sup>. Peer support and the co-production of services are integral to trauma-informed organisations, as relationships are based on mutuality and collaboration<sup>15</sup>. | **Building networks and peer support**<br>Reducing boundaries between professionals and people who use services. This may include more peer support roles, building networks and links between people who use services. Peer support action research is a co-production method that can be used by people with shared experiences to enable greater control over research design and knowledge generation<sup>29</sup>. |
| **Trustworthiness and Transparency**<br>Groups ensure transparency and consistency around how decisions are made, in order to maintain a sense of trust<sup>9</sup>. | **Joint understanding and clarity over roles**<br>**Building and maintaining relationships**<br>Clarity of roles and how the process is led within co-production is important<sup>26</sup>. Building relationships is essential to co-production<sup>6</sup>. It is crucial to understand the temporality and dynamics of trust which means that trust needs to be continuously worked on through a collaborative approach<sup>30</sup>. |
| **Collaboration**<br>Partnership working is vital, with recognition that everyone working together has equal value and importance. Active efforts are made | **Reciprocity and mutuality. Embracing multiple perspectives, experiences and skills. Blurring roles and breaking down boundaries between professionals and people who use services<sup>31, 32</sup>** |
| to identify and reduce power dynamics within groups and between staff and service users to demonstrate how relationships can be healing<sup>9</sup>. | Using the knowledge of all those working together on the research, building on people's assets and the experiences they bring – everyone is of equal importance. Making sure the research team includes all those who can make a contribution, involving diverse stakeholders and being accessible and inclusive<sup>5, 27, 33</sup> |
| **Empowerment and Choice** Groups recognise the resilience and strength of those who have survived trauma. They seek to support people who have survived trauma to develop their own plans and goals, recognising their resilience and ability to heal and promote recovery from trauma. Understand how power differentials may diminish some people's voice and choice. Support shared decision making<sup>9</sup>. | **Sharing power** The work is jointly owned and people work together to achieve a shared understanding, people are working together in more equal relationships<sup>2, 26</sup>. There is a need to consistently reflect on power dynamics and make different forms of power explicit, reflecting on assumptions and usual ways of doing things<sup>26, 34, 35</sup>. |
| **Safety** Groups are in an environment that is not only physically safe from harm but also one in which all group members feel psychologically safe and supported. Interpersonal interactions encourage a sense of safety. Understand what is safe from people's own lived experiences and perspectives<sup>9</sup>. | **Safety** is rarely listed as a core principle of co-production, although time to develop psychological safety with marginalised groups has been highlighted<sup>36</sup>. Participatory research has illustrated the need for safety for both academic and community researchers<sup>37</sup>. Other related issues include risk management and safeguarding. A risk-averse culture has been seen as a barrier to co-production<sup>38</sup>. Risk-averse cultures can be disempowering for peer led co-production methods<sup>18</sup>, which can result in paternalistic professional dependency<sup>39</sup>. Risk awareness and a partnership approach where individual service users are involved in their own safety planning may be more conducive to co-production<sup>19</sup>. |

### Aims and research questions

The aim of Bridging Gaps is to improve access to trauma-informed primary care with women with experience of complex trauma/ needs. This work is ongoing. In this article, we focus on how we used co-production approaches to develop the project and reflect on how we learnt to tailor these to support people who have experienced complex trauma. Our improvement work with general practices will be reported separately. In this article we investigate the following question:

- When working with people who have experienced multiple traumas, how do co-production approaches need to be developed to ensure safe, collaborative and effective working relationships?

### Development of the co-production group

As a general practitioner (GP), co-author LP has delivered a once-a-week outreach clinic in the drop-in centre of support charity, One25, for five years. The outreach clinic is an attempt to provide more accessible healthcare to a highly marginalised group within a trusted community space alongside services delivered by One25 but is unable to offer the full spectrum of mainstream primary healthcare and only operates one day a week. The women that One25 support have experienced complex trauma and face numerous adverse circumstances such as addiction, mental health issues, homelessness, trafficking, domestic violence, sexual exploitation, having children removed from their care, and street sex work. All the women One25 works with have experienced trauma, and it offers specialist services alongside an ethos of non-judgemental, unconditional love. Through her clinical work and conversations with staff and women attending the One25 drop-in centre, LP identified that the existing mainstream primary care system was largely not accessible to the women, despite high levels of clinical need. It was hoped that by bringing the right experts together (GPs, women with lived experience, One25 staff, researchers) better solutions could be developed. This led to the creation of Bridging Gaps.

Co-production meetings were held initially weekly then every two weeks and took place in well-known community spaces that One25 already used to provide services. Participants were offered shopping vouchers as a thank you for their time and contributions following NIHR guidance^40^. The initial goal of these meetings was to discuss the ways in which women with lived experience of trauma could have their needs better met by primary healthcare. As the project developed, the group compiled some information about the project to encourage new members to join (Figure 1). During initial conversations about the project, it was stressed that participation or the choice not to participate would have no bearing on healthcare received and existing support from One25 would continue.

**Figure 1.**
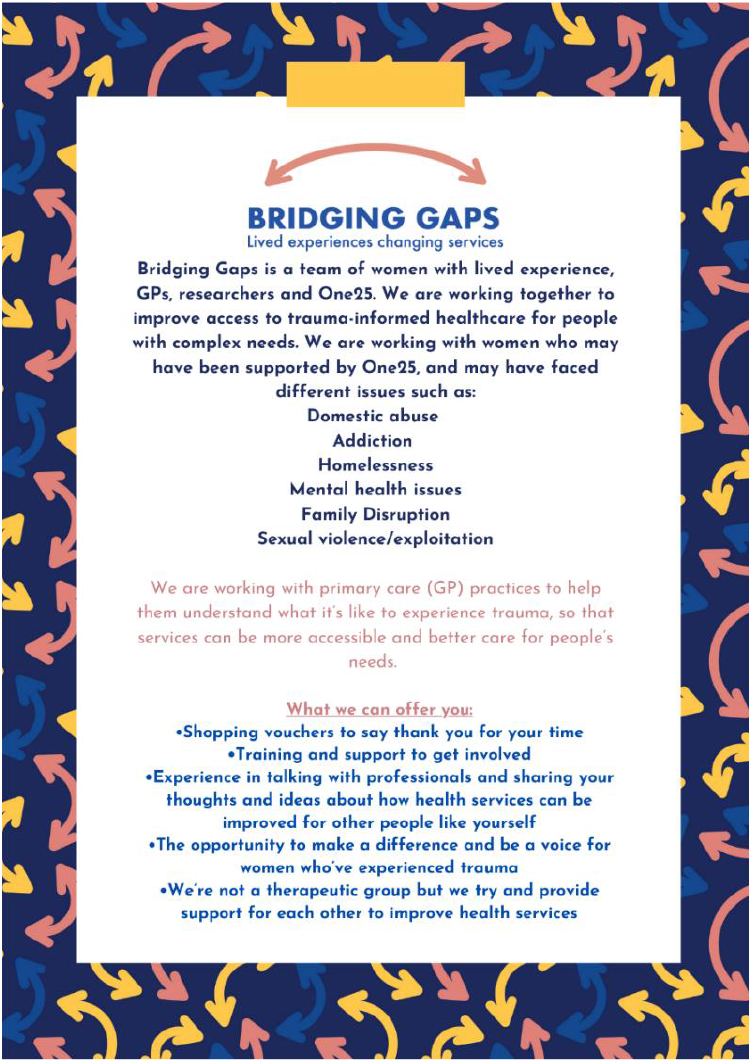
Eligibility criteria and recruitment for Bridging Gaps. Design by Ellie Shipman.

Meetings were attended by at least two professionals (GPs, researchers or staff from One25) to provide assistance e.g. if someone should become distressed and need to be supported within or outside the meeting. Over the period of operation (April 2019 – to present) the core co-production group who met fortnightly has included a total of twenty-nine women with lived experience of trauma, four researchers, two academic GPs, four GP trainees, and four One25 staff members, all of whom were women. Due to the longevity of the project, there has been significant variation in membership of the group. At any one time there have typically been two to six women with lived experience, one to two academic GPs/ GP trainees, one to two researchers and one staff member from One25 attending the groups. Group meetings happened on a fortnightly basis where possible, providing a sense of routine and continuity. Initial recruitment of women with lived experience of trauma took place in collaboration with One25, with LP and One25 staff directly approaching women who may have an interest in the project. Some of the activities of Bridging Gaps over the four years of the project are outlined in Table 2.

**Table 2.**
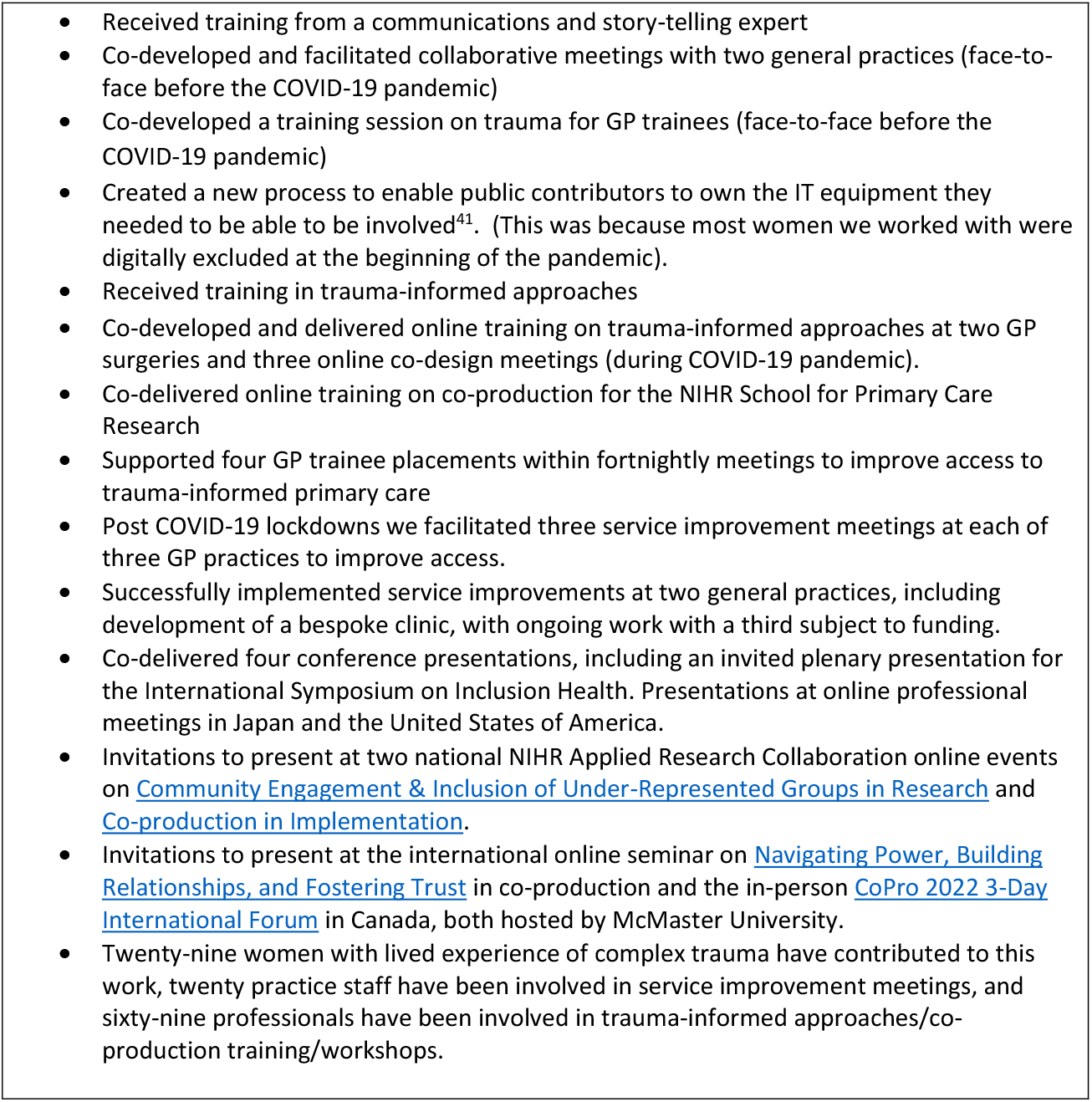
Bridging Gaps activities

## Methods

Methods used for this article were based on participatory action research and co-operative inquiry, where research is developed with people rather than on them^41^. Women with lived experience were much more motivated by practice and the possibility of change, than by theory or research. Where they had been involved in research before, they often saw little change following this. The Bridging Gaps group were focussed on “building participants’ capacity to critique and question current arrangements, and innovate in the development of social practices”^17^(p.86), aiming to improve access to trauma-informed primary care.

### Ethics approval

This research was granted approval by the University of Bristol Faculty of Health Sciences Research Ethics Committee, references 93802 and 110882.

### Data collection

In traditional research the roles of researchers and participants are mutually exclusive. Here all partners and women with lived experience contributed to the design and management of the work, and all were co-researchers and co-participants (for example, researchers were also interviewed by more distant colleagues)^41^. To understand, reflect on and document our approach to co-production, we conducted the following data collection and analysis.

### Reflective notes

At the beginning of the project an academic GP and researcher made reflective notes on fourteen meetings, including project meetings (n=11) with the women, one initial preparatory meeting with GPs not involved in Bridging Gaps and a member of the CCG, one storytelling workshop and one group visit to funders. Further reflective notes on their involvement in the project were made by a researcher, and the four GP trainees involved in the project. Evaluation reports to funders included reflective questions on progress and included input from all members of the co-production group. Whilst we have not quoted from these reflective notes, they provided a helpful record of project development.

### Observation

Observations were conducted of three co-production group meetings and one storytelling workshop, by researchers (HM, MF and HC) who were not at the time involved in group facilitation. Each observation was conducted by two researchers simultaneously with allocated time for discussion and comparison of field notes before these were combined for analysis. The storytelling workshop was one of four sessions led by a professional storyteller. These workshops included various interactive games and activities to help promote bonding as a group and develop skills and confidence in communication and public speaking.

### Semi structured interviews

Twenty-nine women with lived experience have been involved in Bridging Gaps at some time to present (of the twenty-nine members, five have joined since the interviews took place). At the time of the interviews (post COVID lockdowns during summer-autumn 2021), due to life circumstances, reduced possibilities of engagement during lockdowns, and very sadly one death, four women were contactable to be invited to interview. Interviews were also conducted with one researcher, two academic GPs and two One25 staff, all involved in the facilitation/ management of the project. An additional researcher (TS) joined the project to conduct interviews with no prior experience of the meetings. This enabled triangulation between interview analyses conducted by HM and MF who had been involved with the project for over one/ three years respectively with those from TS who had more of an ‘external’ perspective. All interviewees were women, ages ranged from 30-55. Interviews lasted between 19-81 minutes. Separate topic guides were developed for women, partners and researchers (Appendix 1) which were developed and edited by researchers, academic GPs and discussed and agreed with the group. Informed consent (written for face-to-face, verbal audio-recorded for remote) preceded interviews, which were audio recorded, transcribed, checked for accuracy and anonymised.

### Data Analysis

Initially the analytic process involved an inductive approach. HM and HC open coded observation notes with subsequent discussion to develop categories. Reflective notes were analysed in the same ways. Discussion around emerging categories led to the identification of emergent categories relating closely to the six principles of trauma-informed approaches (Table 1) and a framework approach^42^ was adopted in subsequent analysis including interviews. These subsequent analyses were carried out using NVivo 12. Two researchers (TS and HM) double coded two interviews, with subsequent discussion followed by TS coding the remaining interviews.

### Bridging Gaps Lived Experience Input

Bridging Gaps members have prioritised service improvements and achieving change in primary care over involvement in analysis and write up. In-line with trauma-informed collaborative working, this focus was respected and supported while still offering opportunities to review drafts of key recommendations, learning and key points, which were discussed in our fortnightly co-production meetings. One lived experience members who met co-authorship criteria wished to be named as a co-author. Others who would rather not have their real names identified come under the Bridging Gaps group title. Lived experience members have also co-delivered national and international presentations on the project and co-authored a book chapter where the women with lived experience wrote the majority of the words, and academic authors wove together their contributions^43^.

## Results

Results are presented using the overarching framework of the six trauma-informed principles, with additional themes developed inductively and example quotes presented in Tables 3-7.

**Table 3.** Illustrative data for Cultural, Historical and Gender issues

| Themes arising | Data |
| --- | --- |
| Cultural, Historical and Gender issues | **Quote 1:** “Well, what I like about it as well is because it’s women who’ve been involved in – primarily, when it started, it was women who’d been involved in the sex trade, which I have, and I don’t think them women’s voices are heard enough; they’re kind of forgotten about... the sex trade bit of it is kind of hidden and kind of looked down upon and I think it’s really important to get them women’s voices heard and make them feel that they are heard and have a voice in services because they don’t have a voice in that enough.” (Lived experience member 2)<br>**Quote 2:** “Women together and kind of just we have a bit of a giggle, have a cuppa and, you know, it just seems quite easy-going even though we’re talking about quite – it can be quite traumatic, triggering things. Yeah, it feels like a safe space, definitely” (Lived experience member 1)<br>**Quote 3:** “I think women which have got experience at what they’ve gone through and the services that they are possibly using, to have their voices heard and to know that potentially you can make change is totally vital. Because for me, I was silenced for so many years” (Lived experience member 5) |

### Cultural, Historical and Gender issues

Group members chose the term *complex needs* over *severe and multiple disadvantage*^44^ to describe their shared experience of surviving complex trauma. This included experiences of sexual and domestic violence, street-based sex work, trafficking, sexual exploitation alongside homelessness, addiction, having had children removed from their care, and mental health problems (Table 3, Quote 1). The group wanted a female-only environment (Table 3, Quote 2) and we sought to create a non-judgmental, open space where people came together on equal terms, sharing decision-making together and supporting each other: “women can empathise and understand exactly where you are coming from because they’ve experienced similar things themselves” (Lived experience member 5). The group’s aims were developed together with women with complex needs, who had a clear motivation to be heard and change services for others: “we’re a voice for other women being heard” (Lived experience member 9) (see also Table 3, Quote 3). The group reflected how historic experiences of trauma impacted their access to healthcare which could be re-traumatising, and that the needs of women with experience of trauma may differ from those of men. Traumatic experiences impacted the women’s ability to trust and engage with healthcare professionals.

### Peer Support

Initial phases tended to focus on peer support and sharing of experiences. We aimed to enable women to support each other and form meaningful relationships and provided space for difficult and painful stories relating to past interactions with health and social care services to be shared. Early group meetings could involve both laughter and tears, with humour being used as a tool for processing difficult emotions and for group bonding. Women frequently encouraged and supported each other to continue group engagement (Table 4, Quotes 1-2). This highlights the intrinsic value of peer support in enabling women to empower other women. Peer support also extended beyond group meetings, with women informally meeting up separately outside of meetings and checking in with each other when going through difficult times (Table 4, Quote 3). Over time researchers also shared things about their personal lives with the group which helped to engender a sense of equality and community in the group and avoid ‘othering’ (Table 4, Quote 4). Listening to and working with each other also gave researchers a different perspective on their own help-seeking behaviour (Table 4, Quote 5).

**Table 4.** Illustrative data for Peer Support

| Themes arising | Data |
| --- | --- |
| Supporting each other | **Quote 1:** “I’ve not given my kind of story yet but I’ve heard other people’s and seen them do a presentation and it felt supported; it felt like, if it was too much, it |
|  | felt like we'd all support each other...I've definitely got closer to the girls that do the meetings with me, definitely, yeah...I know that if I needed to, I could probably ring them up if I needed to." (Lived experience member 1)<br>**Quote 2:** "The sense of a family and the support from other women, knowing you're not alone, feeling empowered, building your confidence. I could go on, there's so many [laugh], so much, the friendships, not just service level friendships, you really get to know women on a deeper level and I class (Name) as my best friend. I think she changed my life. I could tell her anything." (Lived experience member 9)<br>**Quote 3:** "And so the women at those meetings connect with each other and even if you're not sort of formally kind of creating any peer support mechanisms, those are happening as people kind of walk out meetings together and chat or connect with each other. And I'm aware sort of different women have friendships with each other outside of the group." (Researcher 4) |
| Learning/<br>sharing with<br>each other | **Quote 4:** "I felt like I got a lot closer to [academic GP] and [researcher] and [researcher] confided in me as well and yes, I felt very privileged" (Lived experience member 9).<br>**Quote 5:** "I just remember that actually the time that I did seek help around my own mental health. I was quite inspired by speaking to another member of the team who had talked to me about things she was struggling with.... then I reflected on it myself and so it was the kind of bravery and strength that I see in women with lived experience who do share their stories that did prompt me to say 'okay... maybe actually I need to be brave and share some of my lived experience' and it was a real prompt for me seeking help for myself." (Academic GP 6) |
| Impact of<br>providing<br>support to<br>peers | **Quote 6:** "And then leaving and knowing they're going home alone as well, it stays on your mind. Well it stays on my mind for a bit.....Yes, it's not nice to see that. Like one of the women broke down, yes and it was emotional" (Lived experience member 9)<br>**Quote 7:** "I felt like I had to stand up and be a kind of protector – not protector, that sounds like I'm making myself a martyr and I'm not – but I felt like I had to fight for them a bit more and fight for the kind of – to make sure things are all right for them a bit more" (Lived experience member 2) |

We observed and discussed as a group how the peer support that existed between the women with lived experiences enabled them to challenge the researchers more confidently when there were things they didn’t agree with. Women would feed back that the peer support itself was healing but listening and supporting to each other also had an emotional impact: “it was hard not to be able to take away the pain.” (Lived experience member 9). We became aware that if group members took on too much of this peer support, it could become counterproductive for them and when this happened, we altered our ways of working to try and avoid this (Table 4, Quotes 6-7) (see also empowerment section).

### Trustworthiness and Transparency

Ongoing relationships and trust with One25 staff from the start enabled a safe space to be established in which collaboration could take place. The relationship between the One25 GP (LP) who initially conceived of the project and women with lived experiences of trauma who had been supported by One25 was integral in promoting a sense of trust within the group (Table 5, Quotes 1-2). Trust was also built between women in the group through the sharing of experiences (Table 5, Quote 3). Moreover, the longevity of the project helped in establishing longer term, more trusting relationships (Table 5, Quote 4).

**Table 5.** Illustrative data for Trustworthiness and Transparency

| Themes arising | Data |
| --- | --- |
| Trusting relationships | **Quote 1:** “I think, because what helps with this, is a lot of the women know (Name), she’s a doctor at (One25) she does a clinic there, so a lot of the women would already know her, she would know the women and she understands the sex trade and what all that involves.” (Lived experience member 2)<br>**Quote 2:** “The women have known her for a long, long time and she’s done a lot to serve them really out of (One25). That sort of trust, that’s really hard to get in any other way because that’s just through years of knowing people and showing up for them.” (Academic GP 8)<br>**Quote 3:** “Just the level of trust that all women have really respected and all the women have really shared things as well that they wouldn’t share so it’s then made you feel like wow, like and if you all trust in me then I can trust you. If I’m trusting you, then you can trust me” (Lived experience member 9). |
| Longevity of relationships | **Quote 4:** “Well. I think the longevity of it is really important...women felt that this was a long-term thing, that they weren’t just involved in something for six months and then that ended. That was quite important in terms of women who have experienced a lot of trauma... It was quite important for there to be that time to build up that relationship and trust and then- because that takes ages and once that had been established you had time to deliver things through the project and women to feel, “I am not just being used because of my lived experience” (Support staff 7). |
| Members contributing equally to the project | **Quote 5:** “And I think sometimes...you think, ‘Oh, I’ll just go get the vouchers, put in the bare minimum and I’ve got vouchers.’. I can see it time and time again ... if you’re coming, just doing that, I think it’s not fair on the other women who *are* putting in that kind of work. Sometimes I think a lot of the work can just land on a couple of people and then that’s the – because I know, for me, at times I’ve felt resentment towards other members of the group...because I felt ‘Well, I’m taking on all this ...’” (Lived experience member 2) |
| Transparency around roles | **Quote 6:** “We need to acknowledge the actual existing power dynamics and roles that exist rather than pretending that they are not there and pretending that we are all equal, because I think that backfires and has an opposite effect from what it intended. We have to be really transparent about what everyone’s roles are and the power in that everyone else’s roles and try and mitigate that and try to balance it out a bit. Recognising that we can’t fully flatten it because of the inherent roles that we are playing in the project.” (Support staff 7) |
| Blurring boundaries | **Quote 7:** “Like [support worker] she’s really put her everything into it, like even her personal life. She was all about [Organisation] and the women and like it was more of a friendship. Like she did cross those boundaries with me anyway, I can’t speak for other women” (Lived experience member 9). |

As the project developed a group agreement was discussed and established in a collaborative manner. It was agreed that the group would follow existing One25 guidelines to protect the safety of those within the group e.g. maintaining confidentiality of experiences shared, not being under the influence of substances when at the group. Through the project, some challenges arose with these. Initially when the group first operated on a more drop-in basis, where some women who were in a place of greater instability attended the group, they could talk over others, or kept coming in and out through the meeting. This led to some disruption which was difficult for those who were consistently and meaningfully participating (Table 5, Quote 5). Occasionally, there were instances when women attended the group whilst using substances, potentially threatening the recovery of others. Whilst drug use was not allowed, at times it was challenging to tackle this. Researchers had less experience in identifying signs of drug use and there were sensitivities around accusing or singling women out in the group. As the group established, there were rarely substance use issues arising. Although the aim was to move away from an ‘us’ and ‘them’ approach, professional roles maintained important boundaries that needed to be transparently kept to (Table 5, Quote 6). From a trauma perspective, lack of consistency in boundary enforcement could have a destabilising effect, safety implications and impair trust. In contrast, in other situations, some of the women in the group shared how the crossing of professional boundaries could also support relationships and trust, through the sharing of personal experiences and going beyond boundaries to provide support (Table 5, Quote 7). This relates to the co-production principle of *blurring boundaries*^31, 32^.

Several structural issues complicated our striving to enable more equal relationships. Researchers and support staff held organisational positions which meant that they were ultimately responsible for elements such as budgets, upholding organisational policies and procedures (Table 5, Quote 6). However, when these issues were discussed openly within the group, a greater feeling of trust was fostered. Furthermore, there were opportunities where we could minimise these power imbalances by using transparency. For example, although one of the researchers was the budget holder, decisions were regularly run by the group as a whole on how to spend the budget. These conversations were facilitated by flexible funders, who when asked about budget changes, were able to accommodate the group’s wishes.

### Collaboration

In the early stages of the project there was more flux in group membership as individuals were more or less able to engage depending on what was going on in their life at the time (Table 6, Quote 1).

**Table 6.** Illustrative data for Collaboration

| Theme | Illustrative data |
| --- | --- |
| Group membership fluctuation | **Quote 1:** “I think, going back to when it first started, for some of them women, it was their lifestyle, where they couldn’t keep up the kind of commitment” (Lived experience member 2) |
| Too much email | **Quote 2:** “There was a lot of emails, I think people felt overwhelmed, like it was a job and you were expected to kind of do this, do that. And I know a lot of women wasn’t going to [Researcher] or [Academic GP] but they were coming to me, ‘I can’t handle [it], there’s loads of emails’” (Lived experience member 2). |
| Challenges and disagreements on way forward | **Quote 3:** “Creating a culture and a space where challenge can happen and conflict can happen and be resolved. That is to me the really interesting to learn about.” (Support staff 7)<br>**Quote 4:** “We haven’t had no disagreements as such. We have debates and I’ve walked off and I’ve gone away frustrated which I’m sure some of the GPs and that who are involved and (Name) have gone away frustrated from the way I’ve reacted sometimes but, as a group as a whole, we haven’t had no massive disagreements or falling outs. We’ve managed to come to – once I get over the frustration – we manage to come, we do come to an agreement and kind of work things out. Yeah. And I know everyone’s thing is to make sure the women are safe and in a safe space, to be able to speak about things ...” (Lived experience member 2)<br>**Quote 5:** “I think it’s down to personality. Some women feel more confident to speak up. Other women are a little bit more reserved and perhaps don’t say as much... For me personally, I wasn’t that vocal, I think. I was quite happy to just go along with it ... I think the thing is ... if there were things I possibly didn’t agree with or wanted to say, I knew that I could in confidence either call [Academic GP] or [Researcher] you know, so yeah, I did feel that I could air it with them.” (Lived experience member 5) |

The lead academic GP reflected that this led to her holding group plans and newer members inheriting plans made by others who had left, which may have led to the perception that they were *her* plans as she was the one re-stating them.

Communication was essential to building collaborative relations. Before the COVID-19 pandemic all meetings were face-to-face and one funder required that the group attend training days to share learning with others. The car journey to these training days allowed an opportunity to deepen the relationships in the group. The neutral space of the car and no agenda for conversation may have reduced existing power dynamics, fostering openness and relationship-building. In contrast, the COVID-19 pandemic had a significant impact on our ability to collaborate by preventing meetings in person. We transferred to phone conferencing/ online meetings and not all the women had access to e-mail/ online conferencing initially, until researchers were able to provide IT equipment. When group members did have access to e-mail, people often did not want long emails as it could be too much information (Table 6, Quote 2). These challenges in communication then also had an impact on the relationships between people, where misunderstandings or differences of perspective may have been harder to air. Both support staff and researchers reflected on the substantial need for ongoing relationship maintenance which could be fragile and had potential to be broken. Existing hierarchies may have influenced the women’s freedom to dissent to ideas suggested by the professionals in the team (Table 6, Quotes 3-5), which we discuss further in the empowerment section below.

### Empowerment and Choice

The project evolved from early phases where women spoke about their own personal experiences of trauma to a more focused problem-solving approach when collaborating with local general practices. Through initial group meetings, storytelling workshops and meetings with GP colleagues/ trainees, we developed a series of activities to build confidence in interacting with professionals.

Storytelling workshops were run by an external facilitator, and many of the exercises promoted empowerment and choice. The story telling facilitator provided guiding rules of no self-criticism, creating a safe space in which all group members were “doing the same silly exercises” and where all were free to make mistakes and develop new skills (Table 7, Quotes 1-3). After two workshops with general practices, the group themselves undertook training in trauma-informed approaches, to have a greater technical understanding of issues of trauma. Members of the group varied in their perspectives over whether this was helpful or had the potential to be triggering (Table 7, Quotes 4-5). As we developed the healthcare professionals’ training, women shared their experiences to illustrate how trauma affects people when accessing general practice. Again, the group had differing perceptions on how sharing experiences felt, but at times it could be triggering (Table 7, Quotes 6-7). The combination of sharing experiences and then doing this as part of an online training event with GPs (due to COVID-19) could be difficult, especially when some professionals had their cameras off: “so you couldn’t even see their faces” (Lived experience member 9). The difficulties of interaction online made it difficult for the group to know they were being listened to.

**Table 7.** Illustrative data for Empowerment and Choice

| Theme | Illustrative data |
| --- | --- |
| Storytelling workshops | **Quote 1:** “The work with [Storytelling facilitator] felt like it really brought lots of relationships closer as well... It was women with lived experience, GPs, researchers, all doing the same silly exercises... yeah, it was a real leveller, and it was enjoyable” (Academic GP 6)<br>**Quote 2:** “I think it really helped to do, is build a team, like give that team feeling? And what I liked about it as well – because we had trainee GPs involved – they took part in it as well so it wasn’t ‘Right, we’re the service users over here and you’re the staff.’. That felt very – we was just all on the same level. And I think it’s good it solidified us a group and kind of made us... and just to see the women chatting in a group. Some of them have never even sat in a group and spoke.” (Lived experience member 2).<br>**Quote 3:** “Loved them. Every single one from day one, she’s [storyteller] wicked, amazing. So I think mainly that was what got my confidence up I think, yes... I boob bounced [Researcher] [laugh]. That was wicked that was. You know what I look back on them and every time it makes me smile. Yes, those were straight bonding sessions.” (Lived experience member 9) |
| Training in trauma-informed approaches | **Quote 4:** “It was great.... So a lot of the stuff that we were taught ... I’d already done a lot of it anyway doing the other training, but obviously some of the women coming through hadn’t done any of the stuff, so for them it was a really important thing for them to do.” (Lived experience member 5)<br>**Quote 5:** “I think because we’ve all experienced trauma, to then do that training and try and keep our head focused on actually what we’re just trying to do ... make it easier for women to access primary health care... we just got consumed by this trauma feeling ... it felt like a slog at times.... But it wasn’t no-one’s fault; I think we just got carried away with the trauma side” (Lived experience member 2) |
| Integrating difficult experiences through sharing | **Quote 6:** “So normally would have said talking about my past traumas but that’s actually been like a self-counselling for me to be honest because before I couldn’t talk about it without getting upset or being angry or just feeling loads of emotions that I guess now I can talk about it and I feel fine. It doesn’t bother me in the slightest so it’s kind of been – so I would have said that at the beginning but it’s not that now.... I feel like I can talk about it now without crying or getting angry or going home and feeling, yes, like feeling depressed.” (Lived experience member 9) |
|  | **Quote 7:** “Obviously when you’re telling your own personal story, it can be a bit emotional because it’s not conversations you have on a regular basis. I mean, like me personally, I don’t talk about my trauma that I’ve gone through in the past, and then when you do talk about it, initially I found it quite sort of triggering for me because obviously it brings back a lot of very hurtful memories, but it’s trying to sort of detach yourself from it. I know that I’m in a better place from the fact that I’m not going through any violence.” (Lived experience member 5) |
| Feeling unable to say ‘no’-unrecognised power dynamics in the room | **Quote 8:** “When you’re in recovery – I know I’ve done it when I first got clean and people would say, ‘Oh, can you do that?’, I’d be, ‘Yeah.’, I just wanted to do some[thing].... We’d be in Bridging Gaps and everybody would be like, ‘Yeah.’, and then we’d walk out and say, ‘Do you know, I don’t want to do that’..... So then I’d have to go and say, ‘Look, they’re not really happy with that.’, and they’d be [saying] ‘But they said ‘yeah’.’, and I’d be [saying] ‘Look, you don’t understand. People will say ‘yeah’ because they wanna please you and they’re excited to be part of something and to have their voices heard but’ – and it’s not just on the staff, it’s on the women as well and that’s what I say to them, ‘If it doesn’t feel right, you’re allowed to say no and no one’s going to say, ‘Right, get out’, so I think it was from both sides as well, but if that understanding was there that that’s – and they did, I explained it and we all got through it - but I think there wasn’t that kind of real understanding of how service users can behave.” (Lived experience member 2)<br>**Quote 9:** “I think you need to be working together for a really long time and there needs to be significant trust to say, ‘I don’t really like this’, or ‘let’s go in a different direction.’” (Academic GP 8)<br>**Quote 10:** “Yes a lot of people are like yes people and just like to agree ‘cause they don’t want to hurt your feelings or they don’t wanna be challenged on anything or debated on anything. I don’t know what it is, it could be anything but I feel like people just say yes ‘cause it’s the easier option so it’s always better to ask people first what they think” (Lived experience member 9). |
| Being empowered to choose what to share and when | **Quote 11** “Sometimes I feel I have to overly prove that I’m valid to be there, kind of thing. I’ve got this to prove, you know? So then I’ll overshare, I’ll say, ‘Well, I’ve gone through this.’... I was speaking to [Academic GP] recently and she said she doesn’t go nowhere and say, ‘Well, I’ve done this degree.’, she just goes in, and that kind of did flip a switch ...really it’s, ‘What’s going to be helpful to *you* emotionally as well as to the project?’ and you don’t have to divulge *everything* to make change! You don’t have to prove that, ‘I’ve been through this and this and this.’” (Lived experience member 2) |
| Creating change | **Quote 12:** “You feel like you’ve got, you’ve made a difference. Like got a little bit of purpose as well... you feel like you’re becoming something and that’s a good feeling rather than just sat and [sigh] what have I done today... Yes it helps you...it envisions you to do other things as well. Like you leave there and you can get things done like that you haven’t been able to get done without coming here first... when I leave the meeting, it gives me motivation to do things as well.” (Lived experience member 9) |

These experiences, combined with feedback from our partner One25 and the group, taught researchers an important lesson. Whilst people might initially agree to something, on a later check-in people might change their minds. There were unrecognised power dynamics operating where people might say yes to something in a meeting, when really they want to say no, or might change their minds at a later date. We found that when people agreed to propositions “yes” might not always mean yes and on reflection people might decide differently. An initial “yes” could cover disagreement, uncertainty and fear, that rose to the surface at later dates (Table 7, Quote 8-10). These dynamics could contribute toward misunderstandings or feeling let down or frustrated.

The combination of difficulties in communication over email, the inability to meet face-to-face during the pandemic and unseen power dynamics culminated in a significant turning point for the project. As soon as we were able to shift back to face-to-face meetings after COVID-19 lockdowns we reviewed the training model and removed any needs for the group to share any experience, unless they so wished to informally through general conversation. We found that working in a more trauma-informed way was about empowering people to get involved **and** having the choice to not get involved, moving away from people recounting experiences unless they choose to in the moment (Table 7, Quote 11). As we have progressed through the project and made changes within general practices this has motivated group members to make changes in other aspects of their life (Table 7, Quote 12).

### Safety

Having One25 support staff present within group meetings was essential to provide expertise and continuity, providing guidance to researchers and enabling support for the women if needed, including debriefs and space to discuss any issues or trauma that may have been triggered.

Facilitators were responsible for initiating a check-in and a check-out at each meeting. This involved everyone sharing in turn how they are feeling in the moment, so that support could be offered where needed. In addition, support staff were aware of other issues that may be happening in group members lives and how that might impact their participation (Table 8, Quote 1). There were varying levels of experience in working with people who’ve experienced trauma, with differences according to professional roles, training, knowledge and skills around safety (Table 8, Quotes 2-3). Researchers identified the need for funders to be more aware of the multiple skills, resources and time that this work takes (Table 8, Quote 4).

**Table 8.** Illustrative data for Safety

| Theme | Illustrative data |
| --- | --- |
| Different staff training, skills and knowledge | **Quote 1:** "I think it was difficult in the sense that we don't know people's backgrounds, and that's not appropriate for us to know, but there might be things that are coming up that we're not aware of, or issues that might potentially trigger, or there might be a lot going on in the background of someone's life that we might not be aware of. And that might affect people's involvement ... how might people experience particular parts of the project." (Researcher 4)<br>**Quote 2:** "I suppose the main thing is about having that understanding of the seriousness of the risk that they are at in their daily lives and understanding that |
|  | they might not want to discuss that but helping them to stay safe really” (Support staff 3)<br>**Quote 3:** “As lovely as (Researcher) and (Researcher) are, they don’t have that understanding of that lifestyle and just the chaos of that lifestyle and the up and down of that whole...”. (Lived experience member 2) |
| Resources to support the work | **Quote 4:** “I think sometimes funders and systems are encouraging us to do this kind of thing, but, actually, the difficulties of doing it are quite extensive and probably not accounted for sufficiently, including researchers’ own skills and time and resource to do this.” (Researcher 4) |
| Remote working (during COVID-19 lockdowns) | **Quote 5:** “When we’re all together doing it face to face, we’re very supportive of each other, we can support each other, we all kind of know what each other’s feeling and going through but when you’re kind of stuck at home on Zoom and then you’re sharing all this stuff and then you’re stuck with yourself and all the stuff.” (Lived experience member 2)<br>**Quote 6:** “I think when we were able to meet [online] during lockdowns, that actually felt quite important because there was something there that people could kind of access, but then that also has become quite difficult at times in terms of ensuring safeguarding and confidentiality, if other people are in the house and shared spaces, and not being able to guarantee and know who is in a space, which made it more difficult to kind of operate through lockdowns as we as progressed.” (Researcher 4) |
| Differing perspectives on safety and safeguarding | **Quote 7:** “On safety, we have to consider safety of people and we are responsible for everyone’s safety and so there are just some things that we have to dictate. That’s just how it has to be” (Support staff 7).<br>**Quote 8:** “When we were on Zoom there were some things around actually there might be a perpetrator there in the background..... I think...we were a bit concerned at the beginning about the team’s understanding of safeguarding and actually the fact that women come across as really presentable but actually they have got all these things going on in their background and that that needed to be acknowledged.” (Support staff 3)<br>**Quote 9:** “We were discussing about putting some safeguarding information onto an application form and it immediately evoked a response and she explained that response really clearly as to why even the terminology or any illusions to safeguarding was something that was quite traumatic and something that many women responded badly to who had had bad experiences under the banner of safeguarding” (Academic GP 6) |
| External supervision | **Quote 10:** “I was a bit like, ‘No, I think we really do need clinical supervision’, and so then, fortunately, kind of linked in to get some... got an external person involved. So that was great, and I think that was really important. We’ve now funded, kept into the funding mix that there is that” (Researcher 4). |

When the pandemic and lockdowns began, the group first moved to a free phone-in conferencing system as not all members had access to Wi-Fi or IT equipment to access online conferencing. It became hard to avoid talking over each other, and once we had funders/ IT agreement to buy mobile tablets for the group to own, we moved to online conferencing systems. Remote working led to challenges due to the loss of peer support from in-person meetings “you’re just stuck at home with it” (Lived experience member 2) and some safety concerns (Table 8, Quotes 5-6). When joining online meetings, finding confidential space in your own home is not always possible for women in abusive relationships, and homelessness often means a lack of privacy and safe space. There were varying opinions as to who might be best equipped to make decisions relating to the safety of group members. Support staff felt that they held responsibility for supporting group members’ safety (Table 8, Quotes 7-8), and others questioned how to empower individuals and offer them choices, whilst ensuring the safety of all group members. We resolved this situation through individual safety plans and asked some of the women to join the meetings from a community centre space to guarantee confidentiality for the rest of the group (and paused the project completely when community centres were shut in early 2021). These issues that we grappled with were reflective of wider tensions between safety and empowerment, that the women discussed in the group, where in the past professional actions described as safeguarding had become disempowering or traumatic for the person concerned (Table 8, Quote 9). Both support staff and researchers identified a need for external clinical supervision to help manage some of the more difficult dynamics to support everyone (Table 8, Quote 10).

Another psychological safety issue that arose through our work that was not covered within interviews, was how to manage relational issues when working with new members or general practices where women may have had previous contacts. We developed a process whereby we asked for permission to share first names before a new member joined or discussed who might be at a general practice meeting. This provided some degree of protection but was not infallible as names may not initially be recognised, or triggers might unexpectedly occur at particular places. Debriefs and post-meeting support were essential where unanticipated interactions led to triggering of past trauma.

## Discussion

Whilst co-production processes are to some extent inherently trauma-informed, we identified various areas where additional considerations were necessary. Creation of a safe space is vital. This should include direct consideration of cultural, historical and gender issues which may impact upon group processes. The group should include people already known to and trusted by members who have an understanding of the signs and symptoms relating to trauma. Facilitators should have experience in managing group dynamics and creating supportive environments which empower all individuals to freely express their opinions. Including professionals from a diverse range of backgrounds and having groups based on shared protected characteristics may be of value. Whilst *blurring boundaries* is a key aspect of co-production work, this may need to be approached slightly differently in trauma-informed processes. Transgression of boundaries is a key characteristic of trauma experience, thus maintenance of appropriate and healthy boundaries can be important in creating a safe space for some individuals. Transparency in discussing and agreeing upon group ground rules and how these should be managed is important from the earliest stages of the group. Balancing safety with empowerment and agency was complex. Lived experience members, who had most experience in managing the risks they faced, previously sometimes had difficult experiences under the banner of safeguarding. Researchers had less experience in this field, and support workers felt they held responsibility about holding risk. Further work in this area is needed, incorporating all perspectives.

Whilst consideration of power dynamics is already an important component of co-production, this requires additional attention when working with people who’ve experienced trauma. Those in positions of power need to recognise that people saying “yes” and agreeing to something, might not actually be a representation of people’s true feelings. In her work with women who’ve experienced sexual violence, Ravi^45^ highlights in her *Smile Spectrum* how a smile might be hiding deeper negative feelings and pain. Similarly, we found that when people agreed to ideas, “yes” might not always mean yes, and on reflection, people might want to change their minds. An initial “yes” could cover disagreement, uncertainty, and fear, that rose to the surface at subsequent meetings. We encourage ourselves and others involved in co-production to explore how more safe room can be created on an ongoing basis for disagreement and uncertainty.

Partnership working is key due to the range of skills needed to carry out co-production work with people with lived experience of trauma. For those applying for funding, it is important that adequate resource should be provided for individuals to have flexible access to support between meetings from those with relevant expertise. Additionally, funding applications should consider that time is needed for trust to develop, and maintenance of projects over months and years will be more productive than short-term projects.

Our Recommendations Table 9 (below) is consistent with the experience of other co-produced research with people who have experienced multiple traumas, vulnerability, or complex needs ^5, 6, 25, 46^. Moreover, we add to and highlight existing knowledge in the following ways:

**Table 9.**
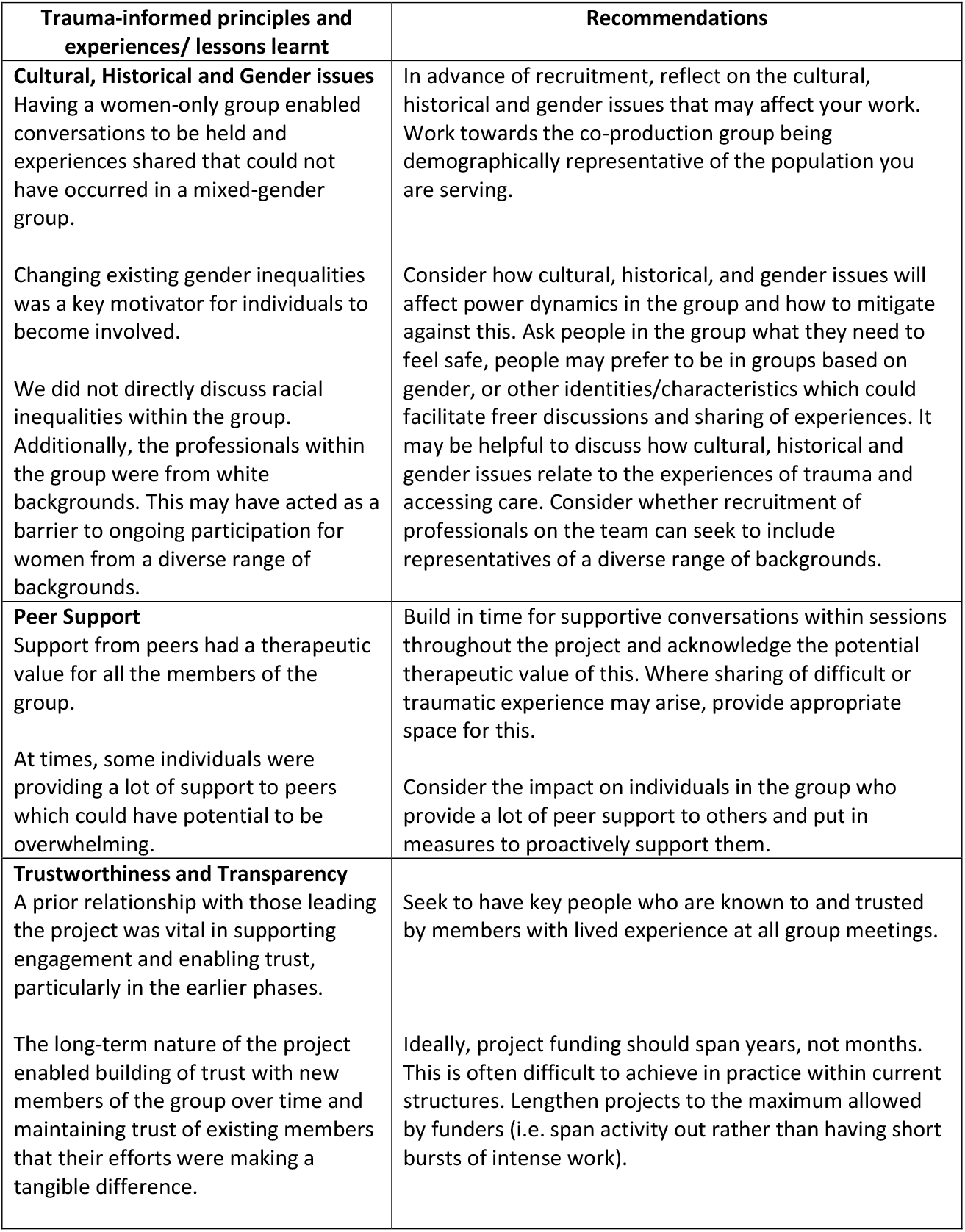
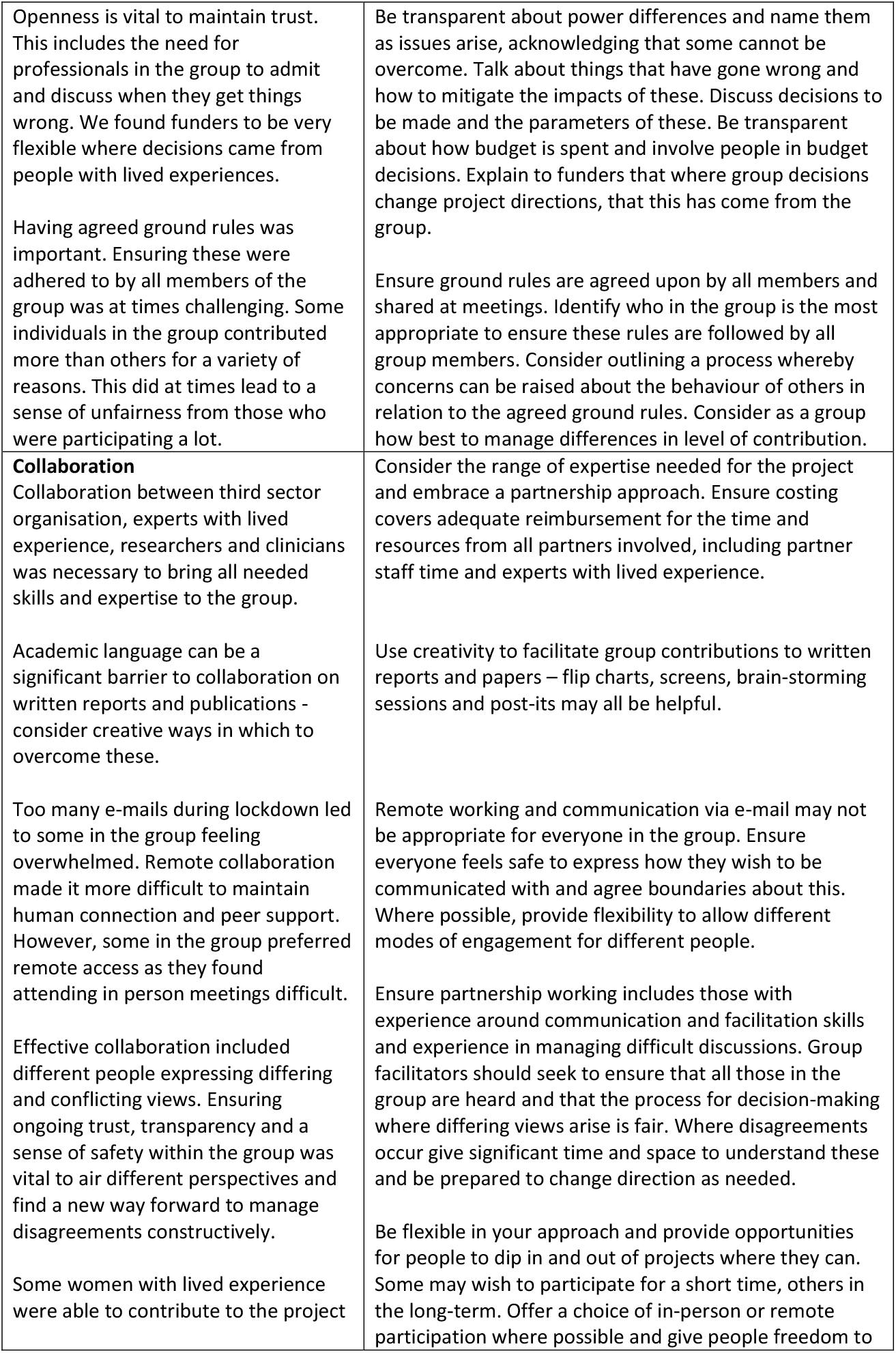
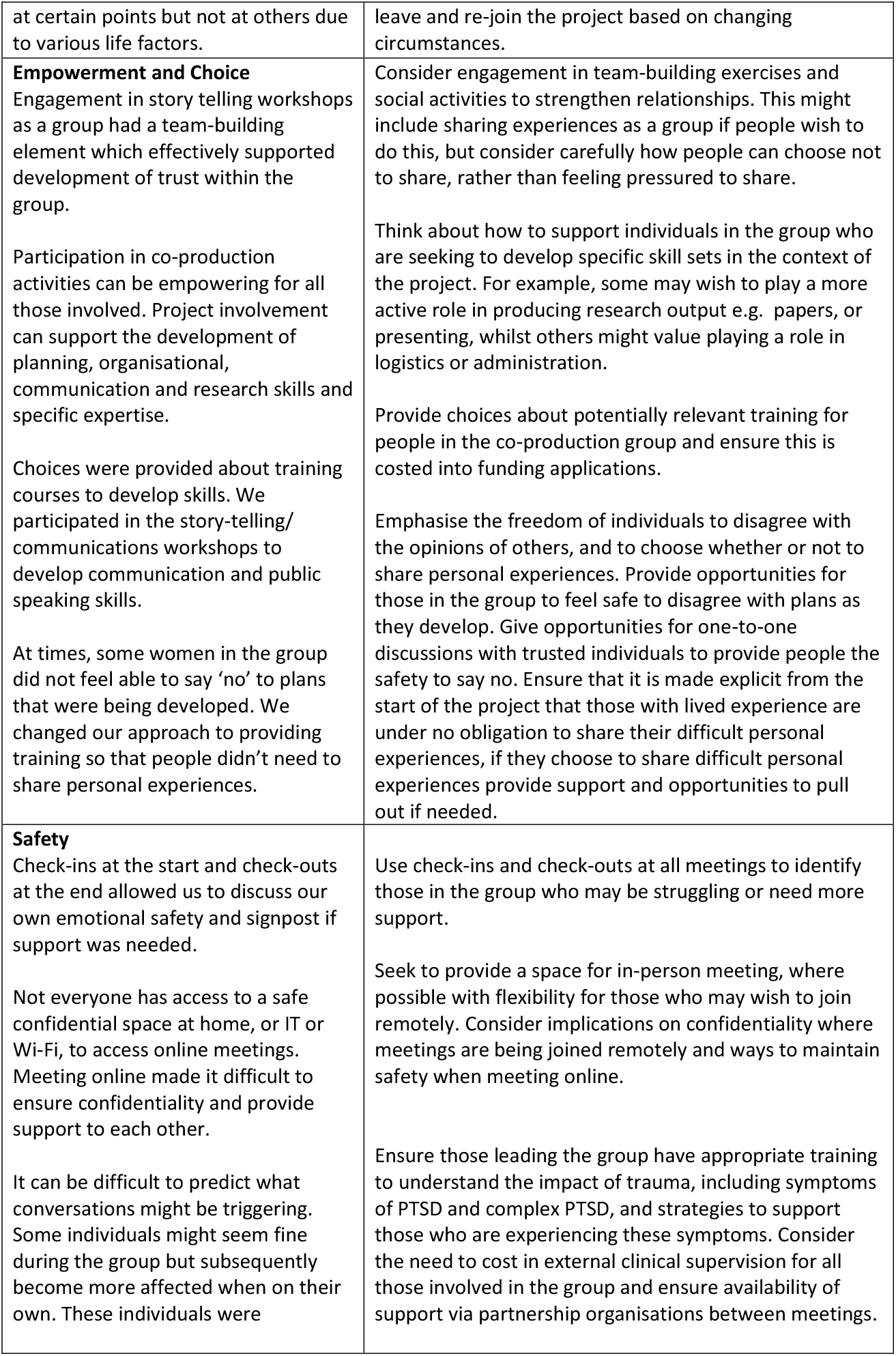
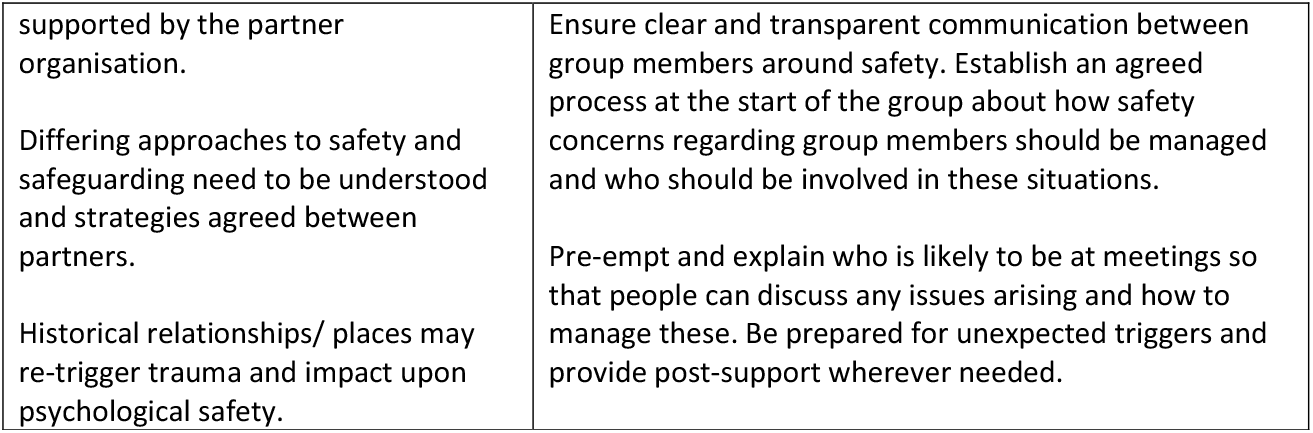
Recommendations table

- The importance of being aware of unseen power dynamics where people may be reticent about expressing dissent or agree to something where they may later change their mind upon reflection.
- Moving away from any emphasis of the sharing of lived experiences to give people full control to choose how, what and when to share, and if they wish to share their expertise and/or experiences. An individual’s lived experience qualifies and enriches their opinion as a valued voice in developing services. That is valid, valuable and enough. They should feel supported to choose to illustrate a point with a personal example if they would like to, or not.
- The challenging dynamics between safety and empowerment. Safeguarding can sometimes mean power and choice is taken away from individuals, and this needs to be managed sensitively and carefully, and recognised and named transparently, explaining the issues and options for moving forward. Furthermore, this needs to be balanced with people’s own expertise in how they manage their own risks^47^.
- Being honest where things go wrong and discussing these openly.
- Greater accounting for the needs of safe and sometimes single gender spaces, or sub-groups based on shared protected characteristics
- The importance of having a safe reflective space for all co-production group members including people with lived experiences, support staff and researchers with external, independent clinical supervision available.
- Reflecting on when you are blurring boundaries and why – what impact is it having and is it helping to provide support and connection, or is it potentially adversely impacting on safety and trust?

Our theoretical contribution is that we explicitly compare and contrast co-production and trauma-informed principles, highlighting differences in safety, how boundaries are managed and maintained and how and when the sharing of experiences may be healing and empowering, or potentially retriggering. This point has significance for the “deeper intellectual shift towards an epistemological position that values ‘knowledge as experience’^48^, and we highlight the caution and care that is needed to ensure that the sharing of experiences is healing and integrative, rather than potentially triggering of past trauma. Whilst sharing of experiences can help to “work through emotional pain”^49^ this process needs to be carefully supported and tailored to the individual at that particular time.

Our project highlights how researchers and practitioners working within these fields can use their social connections and power to help people with lived experience to access funding, resources and knowledge to make changes to services^50^. Through the work there were moments of “transcendent unity”^49^, moving beyond difficulties to create new strength and stability. These insights link with bell hooks^51^ conceptualisation of love, as bringing people together in solidarity, overcoming previous problems^52^. This power through love is not based on personal will, autonomy or sovereignty but on “the desire of well-being of another”, “a way of doing things individually or collaboratively for the wellbeing of others”^52^, love as “connected to the specific values of justice, honesty and generosity”^52^. With this in mind, we share one of women’s artwork with words of her experiences of Bridging Gaps:

> *You are loved*
>
> *You are wise*
>
> *You are a strong independent woman*
>
> *You are brave*
>
> *You are a beautiful lady*
>
> My experience of Bridging Gaps

### Strengths and limitations

Our study’s strengths include that it uses both a data-led and reflective practice approach to identify recommendations and adaptations for those developing trauma-informed services using co-production. Despite the growing interest in trauma-informed services, there has been little attention given to date on the processes of partnership working between professionals, researchers and people with lived trauma experience as they develop services. Without careful consideration of these processes, there is a danger of partnership working being tokenistic at best, and even perpetuating existing power dynamics^28^. This is unlikely to lead to the transformational systems changes necessary to tackle existing inequalities. Future work could include further co-produced service improvement projects based on our recommendations, with further evaluation.

Our project took place over a number of years, which allowed time for reflections and lessons to be learnt. Interviews included a diverse range of perspectives including those from clinical, research, and voluntary sector backgrounds and those with lived experience of trauma. Analyses were carried out jointly between those who were involved in group processes, and those who were external, enabling triangulation between different viewpoints when analysing the data. Observational and reflective data were triangulated with interview data. Although the pandemic led to many difficulties in our group processes, it provided some valuable learning on the opportunities and difficulties of remote working for those with lived experience of trauma.

One limitation was the fact that changes in group membership meant that many women who were involved for certain periods of the project could not be interviewed about their experience. Whilst those interviewed suggested that these women dropped out for reasons unrelated to the project it would have been helpful to have their perspectives and reflect on whether any additional support may have enabled them to remain part of the process.

## Conclusion

Our findings provide vital learning points for all those seeking to develop trauma-informed services and an opportunity for further evaluation of our recommendations for practice. The high prevalence of trauma in the general population also makes our findings even more broadly applicable and merit consideration for all those engaging in co-production work.

## Data Availability

Original data are not available due to the potential to compromise people's anonymity and the small number of interviews that were undertaken.

## Acknowledgements

Many thanks to all the women with lived experience of trauma who have been part of this group through its development. Many thanks to all the members of One25 staff that supported this project through its development and gave their time to enable this project to happen. We would like to thank Lesley Wye for all her time and efforts to initiate and support the development of Bridging Gaps. Many thanks to the GP trainees who joined us as part of their placements for their enthusiasm and willingness to get involved. Thanks to all the general practices that we have engaged with, and the work they have done to improve access to trauma-informed primary care.

## Author contributions statement

LP started the Bridging Gaps group with support from MC (and LW – see acknowledgments) and was PI on three grants, co-ordinating the group’s activities and facilitating their interactions with general practices. HM joined Bridging Gaps as a GP trainee and Academic Clinical Fellow, observed sessions and analysed all data and wrote the first draft of this paper. MF joined the group as a researcher and initially observed sessions. She then joined the group and became PI on two grants, co-ordinating the group, its work and its finances. She has supported HM in the development of the writing of this article, including the comparison of co-production and trauma-informed principles. TS conducted interviews with the group, analysed all interview data and contributed to the drafting of this paper and its theoretical analysis. HC observed Bridging Gaps meetings, supported HM in initial data analysis of observations, conducted an interview and contributed to the drafting of this paper and its theoretical analysis. MC was employed by One25 as a support manager and helped facilitate the group and provide support to women with lived experience. She contributed to conversations about the comparison of co-production and trauma-informed principles and the learning of the project. FC is employed by One25 as a support worker and joined the group when MC left One25, contributing to facilitating the group, providing ongoing support, and input into meetings with general practices. JH was academic supervisor for LP and MF. GF was main supervisor of LP. BG is a group of women who have experienced complex trauma and have faced numerous adverse circumstances such as addiction, mental health issues, homelessness, domestic or sexual violence, having children removed from their care, and street sex work. All authors commented on and edited the draft of this paper and contributed to the final manuscript.

